# Redefining ALS: Large-scale proteomic profiling reveals a prolonged pre-diagnostic phase with immune, muscular, metabolic, and brain involvement

**DOI:** 10.1101/2025.08.20.25334061

**Authors:** Jan Homann, Roxanna Korologou-Linden, Vivian Viallon, Sarah Morgan, Valerija Dobricic, Laura Deecke, Julia P. Schessner, Karl Smith-Byrne, Daniel Birtles, Yujia Zhao, Joanne Wuu, Fanny Artaud, Fatema Hajizadah, Jose Maria Huerta, Olena Ohlei, Mikhail Lebedev, P. Martijn Kolijn, Marcela Guevara, Ana Jimenez-Zabala, María José Sánchez, Camino Trobajo-Sanmartín, Sandra M. Colorado-Yohar, Sonia Alonso-Martín, Dafina Petrova, Sabina Sieri, Klaus Berger, Susan Peters, Nick Wareham, Rudolph Kaaks, Ruth C. Travis, Roel C. H. Vermeulen, The Global Neurodegeneration Proteomics Consortium (GNPC), Ioanna Tzoulaki, Alexis Elbaz, Matthias Mann, Carlotta Sacerdote, Giovanna Masala, Verena Katzke, Michael Benatar, Lars Bertram, Lefkos Middleton, Elio Riboli, Marc J. Gunter, Pietro Ferrari, Oliver Robinson, Christina M. Lill

## Abstract

**Background:** Amyotrophic lateral sclerosis (ALS) is a fatal neurodegenerative disorder with a largely unknown duration and pathophysiology of the pre-diagnostic phase, especially for the common non-monogenic form.

**Methods:** We leveraged the European Prospective Investigation into Cancer and Nutrition (EPIC) cohort with up to 30 years of follow-up to identify incident ALS cases across five European countries. Pre-diagnostic plasma samples from initially healthy participants underwent high-throughput proteomic profiling (7,285 protein markers, SomaScan). Cox proportional hazards models based on 4,567 participants (including 172 incident ALS cases) were used to identify protein biomarkers associated with future ALS diagnosis. Top results were indirectly validated in two independent case-control studies of prevalent ALS (n=417 ALS, 852 controls). Functional annotation included cross-disease comparisons, gene set and tissue enrichment testing, organ-specific proteomic clocks, and the application of large-language models (LLM).

**Findings:** Five proteins (SECTM1, CA3, THAP4, KLHL41, SLC26A7) were identified as significant pre-diagnostic ALS biomarkers (FDR=0.05), detectable approximately two decades before diagnosis. Of these, all except SECTM1 were indirectly validated in independent cohorts of prevalent ALS cases, supporting their clinical significance. Additionally, 22 nominally significant (p<0.05) pre-diagnostic biomarkers were FDR-significant in prevalent ALS with consistent effect directions. Cross-disease comparisons with pre-diagnostic Parkinson’s and Alzheimer’s disease suggested a largely specific pre-diagnostic ALS biomarker signature. Gene ontology and tissue enrichment highlighted early involvement of immune, muscle, metabolic, and digestive processes. Furthermore, analyses of proteomic clocks revealed accelerated aging in brain-cognition, immune, and muscle tissues before clinical diagnosis. Druggability and LLM analyses revealed possible therapeutic targets and novel strategies, emphasizing translational relevance.

**Interpretation:** Our study provides first evidence of ultra-early molecular changes in common ALS up to two decades prior to clinical onset, mainly affecting immune, muscle, metabolic, digestive, and cognitive systems. Our study nominates several compelling candidates for risk stratification studies and novel therapeutic targets for early intervention.

**Funding:** Clinical Research in ALS and Related Disorders for Therapeutic Development (CreATe) Consortium, Cure Alzheimer’s Fund, Michael J Fox Foundation, Interdisciplinary Centre for Clinical Research, University Münster

## INTRODUCTION

Amyotrophic lateral sclerosis (ALS) is a devastating neurodegenerative disease, characterized by progressive loss of central and peripheral motor neurons. This neurodegeneration leads to muscle atrophy, rapid functional decline, and usually death within a few years after symptom onset. The disease places substantial physical, emotional, and financial burdens on patients, families, and caregivers^1^.

While 5-10% of patients carry highly penetrant mutations and often show familial aggregation (“familial ALS”), the more common form is non-monogenic and usually does not show familial clustering (“sporadic”). It is likely caused by a combination and interaction of genetic and lifestyle/environmental risk factors. Although several genetic^2^ and environmental/lifestyle^1^ risk factors have been identified, the fundamental pathophysiological mechanisms and their timing underlying ALS remain incompletely understood. This limited insight contributes to the absence of effective disease-modifying therapies; current treatments are mostly symptomatic offering only modest improvements in survival and quality of life^1^.

Unlike Parkinson’s (PD) and Alzheimer’s disease (AD), where pre-diagnostic phases with initial pathophysiological changes and unspecific symptoms may span decades, the timing of this initial phase in ALS is largely unknown, particularly for the common non-monogenic form^3^. This is largely due to the low incidence of ALS (∼2-3 cases per 100,000 individuals annually^1^) and high short-term mortality^4^, which requires very large prospective cohorts with long-term follow-up to investigate this phase. Moreover, the identification of pre-diagnostic markers — such as blood-based biomarkers indicative of risk exposures and/or early pathophysiological changes preceding clinical diagnosis — depends on the availability of carefully stored baseline (i.e., pre-diagnostic) blood samples from such large cohorts. As a result, pre-diagnostic biomarker signatures for ALS remain underexplored to date, especially for the common form: One prospective study including 84 individuals who developed ALS and 161 matched controls found plasma neurofilament light chain (NfL) levels to be elevated already within five years before ALS diagnosis^5^. Furthermore, a study focusing on monogenic ALS found increases in NfL levels in asymptomatic carriers about one year before symptom onset^3,6–8^.

At the same time, high-throughput proteomic technologies such as the aptamer-based SomaScan® (SomaLogic, Inc.) technology offer promising opportunities for the discovery of novel biomarker signatures by enabling simultaneous quantification of thousands of proteins. In this context, blood-based proteomic biomarkers are attracting increasing interest because they are minimally invasive, readily applicable in clinical practice, and have already demonstrated strong predictive performance in other conditions^9–11^, including neurodegenerative diseases like AD^12,13^.

To advance insight into the pre-diagnostic phase of ALS, we leveraged data from the ‘European Prospective Investigation into Cancer and Nutrition’ (EPIC) cohort (n=521,000)^14,15^, which offers 30 years of follow-up, to identify incident ALS cases. This enabled us to establish a well-characterized, population-based case-cohort — hereafter referred to as EPIC4ALS — comprising 4,567 participants, including 172 incident ALS cases^16^. We utilized baseline (pre-diagnostic) plasma samples collected from all initially healthy EPIC4ALS participants to conduct a comprehensive high-resolution proteomic screen for biomarkers predictive of subsequent ALS development using the SomaScan 7K array (∼7,000 protein markers). We tested the most promising pre-diagnostic biomarkers for ‘indirect’ validation in two independent studies of individuals with established ALS. To further contextualize these findings, we performed functional annotation analyses, including the assessment of organ-specific aging in pre-diagnostic ALS, and compared proteomic signatures from pre-diagnostic ALS with those from incident PD and AD. This multifaceted approach allowed us to characterize ultra-early molecular changes and disease-specific signatures in incident ALS, providing new perspectives on disease initiation.

## RESULTS

### EPIC4ALS case-cohort

The EPIC4ALS case-cohort comprises 4,737 participants including a randomly drawn subcohort and 193 ALS cases identified by screening mortality records from 222,613 EPIC participants across five European countries (Spain, Italy, the Netherlands, the UK, Germany) over 30 years of follow-up. Age at death was used as a proxy for age at diagnosis in the Cox proportional hazard regression analysis. In this context, to identify pre-diagnostic biomarkers, all primary analyses were initiated five years after baseline, i.e., five years after blood collection, by excluding 21 ALS cases and 149 subcohort members who died or were censored within this initial window. This yielded 4,567 EPIC4ALS participants (including 172 incident ALS cases; **Figure 1, Supplementary Tables 1**-**2**). These participants had a median age of 52 years at baseline (range: 35–79), 66% were women (62% excluding Utrecht, which recruited only women), and the median follow-up was 18 years. Among incident ALS cases, median time from blood sampling to death was 14 years (range: 5-29). As expected, given our case-cohort setting, incident ALS cases had a higher median age at baseline (58 vs. 52 years) and a greater proportion of men (47% vs. 38%, excluding Utrecht) compared to non-cases (please note that these differences are appropriately accounted for by stratification for age and sex in our Cox models). Additionally, ALS cases were less likely to be overweight (BMI ≥25: 49% vs. 61%) and less likely to be current smokers (17% vs. 24%; **Supplementary Table 1**).

**Figure 1.**
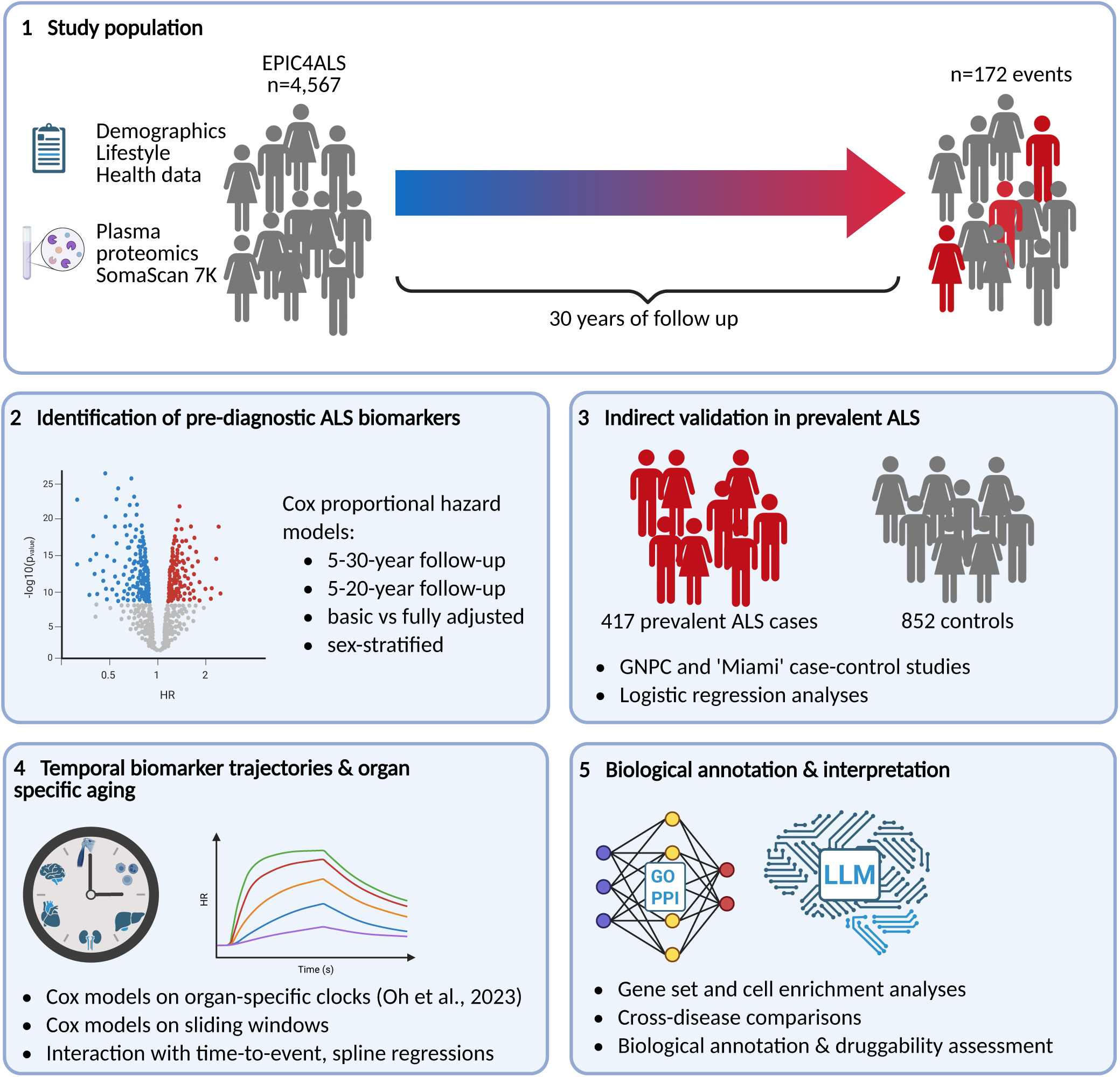
Visual summary of study design. <u>Legend.</u> In this study, plasma proteomics from 4,567 participants (172 incident amyotrophic lateral sclerosis [ALS] cases) from the ‘European Prospective Investigation into Cancer and Nutrition for amyotrophic lateral sclerosis’ case-cohort (EPIC4ALS) were generated to identify pre-diagnostic protein biomarkers of ALS. Using the SomaScan 7K platform, 7,285 aptamers were analyzed across up to 30 years of follow up. Identified biomarkers were indirectly validated in two large datasets (Global Neurodegenerative Disease Consortium [GNPC] and the ‘Miami’ dataset by Dergai et al.^31^) including 417 prevalent ALS cases and 852 controls. Accelerated biological aging was investigated using organ-specific proteomic clocks^20^. Further functional characterization included gene-set and cell-type enrichment, protein-protein network analyses, the application of large-language models (LLM) models, as well as cross-trait analyses with Parkinson’s (PD) and Alzheimer’s disease (AD).

### Association analyses in the EPIC4ALS discovery dataset

To identify proteins associated with a future ALS diagnosis, we profiled 6,381 proteins (using 7,285 aptamers) in plasma samples of 4,567 EPIC4ALS participants, including 172 incident ALS cases. Cox regression analyses, stratified by baseline age (in 5-year intervals), sex, and center, identified only the SECTM1 protein as significantly associated with ALS risk over 5-30 years of follow-up (hazard ratio [HR]=1.35 per standard deviation [SD] increase in log10 protein levels, p=1.38×10^−6^) at a false-discovery rate (FDR) control of 0.05. Restricting analyses to 5-20 years of follow-up, five proteins were FDR-significantly associated with ALS risk: SECTM1, CA3, THAP4, KLHL41, and SLC26A7 (all HR>1.30, p≤2.77×10^−5^; **Table 1, Figure 2A, Supplementary Figure 1**). Analysis of Schoenfeld residuals for all five biomarkers during 5-20 years follow-up showed no deviation from the proportional hazards assumption (data not shown). Additional adjustment for demographic and lifestyle factors had minimal impact, with effect size estimates for the top biomarkers remaining stable (**Supplementary Figure 2, Supplementary Table 3**). Sex-stratified analyses revealed stronger associations for CA3, THAP4, and SLC26A7 in men than in women (ratio of relative risks >1.30, all p<0.05 for sex interaction; **Figure 2B, Supplementary Table 4**). Sensitivity analyses excluding outliers instead of capping (see Methods) yielded similar effect size estimates, supporting the robustness of our top results (**Supplementary Figure 2, Supplementary Table 5**). Three biomarkers (KLHL41, CA3, SLC26A7) were moderately inter-correlated (r=0.30–0.54), suggesting shared regulation or pathways, whereas SECTM1 and THAP4 were largely uncorrelated with the others (**Supplementary Figure 3, Supplementary Table 6**).

**Table 1.** Plasma protein biomarkers significantly associated with future onset of amyotrophic lateral sclerosis.

|  | EPIC4ALS, incident |  | GNPC, prevalent |  | 'Miami', prevalent |  |
| --- | --- | --- | --- | --- | --- | --- |
| Protein | HR (95% CI) | p | OR (95% CI) | p | logFC | q |
| ***FDR-significant pre-diagnostic biomarkers*** |  |  |  |  |  |  |
| SECTM1 | 1.42 (1.24-1.62) | **1.61E-07** | 0.97 (0.75-1.24) | 7.79E-01 | -0.03 | 5.00E-01 |
| CA3 | 1.43 (1.24-1.66) | **9.75E-07** | 3.33 (2.48-4.45) | **6.75E-16** | 1.21 | **6.04E-35** |
| THAP4 | 1.41 (1.22-1.64) | **6.37E-06** | 1.89 (1.39-2.56) | **4.92E-05** | 0.13 | 6.80E-02 |
| KLHL41 | 1.32 (1.16-1.49) | **1.44E-05** | 3.06 (2.32-4.03) | **2.70E-15** | 0.79 | **6.90E-21** |
| SLC26A7 | 1.32 (1.16-1.51) | **2.77E-05** | 3.27 (2.18-4.91) | **9.47E-09** | 0.11 | **1.20E-03** |
| ***Suggestive pre-diagnostic biomarkers*** |  |  |  |  |  |  |
| DUSP29 | 1.28 (1.12-1.45) | 2.15E-04 | 4.46 (2.85-6.96) | **5.46E-11** | 0.30 | **1.27E-07** |
| ARL5B | 1.26 (1.09-1.45) | 1.34E-03 | 1.55 (1.17-2.05) | 2.28E-03 | 0.11 | **3.80E-02** |
| CD36 | 0.76 (0.64-0.90) | 1.52E-03 | 0.71 (0.54-0.93) | 1.29E-02 | -0.28 | **1.40E-03** |
| PDLIM3 | 1.26 (1.09-1.45) | 1.68E-03 | 6.40 (4.25-9.62) | **5.65E-19** | 1.57 | **1.73E-44** |
| CST3 | 1.28 (1.09-1.51) | 2.41E-03 | 1.31 (1.01-1.70) | 4.36E-02 | 0.12 | **7.40E-03** |
| DCT | 0.72 (0.58-0.89) | 2.68E-03 | 0.75 (0.56-1.00) | 4.72E-02 | -0.18 | **3.00E-02** |
| ATF5 | 1.24 (1.08-1.43) | 2.81E-03 | 2.66 (1.83-3.86) | **2.59E-07** | 0.08 | 4.70E-01 |
| MYL6B | 1.23 (1.07-1.42) | 4.70E-03 | 3.84 (2.58-5.72) | **3.64E-11** | 0.35 | **4.83E-11** |
| HSPB6 | 1.39 (1.10-1.74) | 5.33E-03 | 3.91 (2.70-5.65) | **4.31E-13** | 0.58 | **6.90E-21** |
| MYL3 | 1.27 (1.07-1.50) | 5.34E-03 | 2.22 (1.66-2.96) | **6.61E-08** | 0.55 | **3.36E-08** |
| ACYP2 | 1.43 (1.10-1.86) | 8.47E-03 | 1.87 (1.35-2.60) | **1.92E-04** | 0.06 | 4.10E-01 |
| CAPN3 | 1.17 (1.03-1.33) | 1.40E-02 | 4.38 (2.84-6.74) | **1.91E-11** | 0.56 | **8.19E-13** |
| DCTN2 | 1.24 (1.05-1.48) | 1.40E-02 | 1.21 (0.94-1.56) | 1.32E-01 | 0.08 | **1.00E-02** |
| CD209_2 | 0.79 (0.64-0.96) | 1.72E-02 | 0.73 (0.57-0.95) | 1.77E-02 | -0.17 | **3.70E-02** |
| CD209_52 | 0.80 (0.67-0.96) | 1.82E-02 | 0.73 (0.59-0.92) | 6.24E-03 | -0.31 | **7.07E-05** |
| MYBPC1 | 1.19 (1.02-1.40) | 2.80E-02 | 3.49 (2.56-4.74) | **1.83E-15** | 0.85 | **2.28E-14** |
| MYOM2 | 1.20 (1.02-1.42) | 2.83E-02 | 3.66 (2.64-5.07) | **5.53E-15** | 1.19 | **2.11E-20** |
| CKB\|CKM | 1.19 (1.02-1.39) | 3.17E-02 | 2.43 (1.88-3.15) | **1.68E-11** | 0.78 | **1.76E-08** |
| SCN3B | 1.18 (1.01-1.38) | 3.48E-02 | 1.19 (0.82-1.73) | 3.49E-01 | 0.26 | **5.74E-04** |
| ATF6 | 0.81 (0.66-0.99) | 3.63E-02 | 0.90 (0.70-1.15) | 3.89E-01 | -0.20 | **7.07E-06** |
| EHD2 | 1.21 (1.01-1.46) | 4.35E-02 | 1.28 (0.99-1.66) | 5.88E-02 | 0.14 | **2.20E-02** |
| PCDHGA10 | 1.17 (1.00-1.36) | 4.59E-02 | 2.79 (1.94-4.01) | **2.71E-08** | 0.36 | **7.97E-07** |
| MFAP5 | 0.78 (0.61-1.00) | 4.94E-02 | 0.58 (0.42-0.81) | 1.17E-03 | -0.24 | **9.25E-08** |

Legend. This table presents all plasma proteins showing significant (false discovery rate [FDR]=0.05) association with incident amyotrophic lateral sclerosis (ALS) in Cox proportional hazard regressions (upper tier) as well as “suggestive” proteins in EPIC4ALS. The latter are nominally significantly ( $p < 0.05$ ) associated with incident ALS in EPIC4ALS and show FDR-significance in prevalent ALS on a proteome-wide scale in at least one of the two prevalent case-control datasets and with consistent effect directions in all datasets (lower tier). Cox proportional hazard regression analyses were conducted on a follow-up of 5-20 years and stratified for center, sex and 5-year age categories. Note that DUSP29 is also known as DUPD1. GNPC=Global Neurodegenerative Disease Proteomics Consortium. ‘Miami’=data pre-published in Dergai et al.<sup>31</sup>. HR=hazard ratio; OR=odds ratio, logFC=log fold change, CI=95% confidence interval; p=p value, q=FDR-adjusted p value (as provided in <sup>31</sup>).

**Figure 2.** Association results of plasma proteins in the EPIC4ALS case-cohort. <u>Legend.</u> This figure shows association results of protein biomarkers with amyotrophic lateral sclerosis (ALS) based on Prentice-weighted Cox proportional hazard regressions (stratified for age, sex, and center) in the EPIC4ALS cohort. (**A**) Volcano plot of Cox proportional hazard regression analyses across 4,567 EPIC4ALS participants (including 172 incident ALS cases) across 5-20 years of follow-up and stratified for center, sex, and age in 5-year categories. Protein names highlighted in bright blue indicate proteins showing significant association with incident ALS after false-discovery rate (FDR) control (FDR=0.05). (**B**) This figure shows hazard ratios (HR) with 95% confidence intervals for all FDR-significant pre-diagnostic ALS biomarkers upon stratification for sex across 5-20 years of follow-up. Protein names in bold indicate association results that showed significant interaction (p<0.05) by sex. (**C**) Time-varying hazard ratios of top proteins derived from spline regression across the full follow-up of 0-30 years. For these analyses, the 21 incident ALS patients and 149 non-cases who died or were censored within five years of baseline and were excluded from the main analyses were re-included. (**D**) Triangle plot showing an integrated overview of analysis results for proteins across incident and prevalent ALS, cross-disease comparisons, and gene expression and protein levels in relevant target tissues. Proteins showing FDR-significance in the pre-diagnostic EPIC4ALS analyses are listed in the upper tier, proteins showing nominally significant (p<0.05) pre-diagnostic associations and FDR-significance in the prevalent case-control datasets are listed in the lower tier. Triangles pointing upwards indicate a positive association of protein levels with disease risk or status, triangles pointing downwards indicate a corresponding inverse association. (**E**) This figure displays the EPIC4ALS association results of organ-specific proteomic clocks as a measure of organ-specific accelerated biological aging with incident amyotrophic lateral sclerosis (ALS) based on Cox proportional hazard regressions (stratified for age, sex, and center). Organ-specific clocks were calculated as described by Oh et al.^20^. Left: Forest plot of results of pre-diagnostic clocks in EPIC4ALS. Pre-diagnostic organ clocks that surpassed FDR=0.05 are highlighted in color. Note that the ‘organismal’ clock refers to a model that uses the non-organ-specific proteins only, and that the ‘conventional’ clock refers to a model that uses all proteins regardless of organ specificity. Right: Time-varying hazard ratios of top proteins derived from spline regression across the full follow-up period (0-30 years).

### Analyses of time-varying effects

Next, in exploratory analyses, we examined whether and how the strength of the effect size estimates of the top 5 pre-diagnostic proteins varied across the entire follow-up time from the distal pre-diagnostic phase to the proximal phase prior to ALS-related death (prevalent ALS). To this end, we extended the dataset using 4,737 EPIC participants including all 193 ALS cases (i.e., re-including the 21 incident ALS patients and 149 non-cases who died or were censored within five years of baseline). HRs based on 10-year sliding windows suggested comparatively stable HRs for most biomarkers, demonstrating long-term risk associations 25-15 years or even earlier before death, i.e., ∼20-10 years before diagnosis. We observed that HRs of all biomarkers except SECTM1 were slightly larger in time windows closer to ALS-related death (**Supplementary Figure 4**). Accordingly, interaction analyses with a ‘protein level × log time-to-event’ interaction term indicated nominally significant interactions (increasing risk with decreasing time-to-death) for all proteins except SECTM1 (**Supplementary Table 7**). The interaction analysis for SECTM1 showed an inverse (decreasing risk with decreasing time-to-death) but non-significant interaction estimate. Spline-based Cox models to assess potential non-linear relationships demonstrated statistical evidence for a time-varying effect for KLHL41 (p=0.0455), with larger effects in short-term risk settings (**Figure 2C, Supplementary Table 7)**. For SECTM1, CA3, THAP4, and SLC26A7, the spline-based interaction terms were not statistically significant (p>0.11), suggesting that they showed relatively constant performance of predicting ALS risk over the 30-year follow-up period supporting their long-term predictive potential.

While it should be noted that these findings are based on relatively small sample sizes, they globally suggest that biomarker associations with ALS risk can be observed and are relatively stable starting in the time windows of 25-15 years before death (i.e., 20-10 years before diagnosis). This underscores the importance of comprehensive baseline proteomic profiling with longitudinal follow-up to reveal pre-diagnostic biomarker signatures in ALS.

### Indirect validation in prevalent ALS

Due to the lack of sufficiently large pre-diagnostic ALS cohorts with SomaScan data — and since strong effect estimates were seen near or after clinical diagnosis in EPIC4ALS — we used two independent prevalent ALS case-control datasets (417 ALS cases, 852 controls) for indirect validation. These datasets included SomaScan 7K data from the Global Neurodegeneration Proteomics Consortium (GNPC)^17^, which we re-analyzed, and from the University of Miami, with publicly available summary statistics^18^. Notably, all four FDR-significant pre-diagnostic biomarkers except SECTM1 were also significantly associated with prevalent ALS status (with FDR-significance on a proteome-wide scale), with effects in the same direction (**Table 1, Figure 2D**). Of the three biomarkers with sex-specific effect estimates pre-diagnostically, CA3 and THAP4 showed similar differences in GNPC prevalent cases, although the interaction test was not significant (**Supplementary Table 4**).

Of all biomarkers nominally significantly associated with pre-diagnostic ALS but not passing FDR control (n=326), 22 proteins (23 aptamers) showed consistent (proteome-wide) FDR-significant associations with prevalent ALS in both case-control datasets, suggesting genuine — though potentially weaker— associations in pre-diagnostic ALS (“suggestive” biomarkers; **Table 1, Supplementary Table 8**). Among nominally significant pre-diagnostic biomarkers, we found significant enrichment of FDR-significant clinical ALS biomarkers (odds ratio [OR]=4.06, p=3.63×10^−6^ [GNPC]; OR=2.27, p=8.35×10^−5^ [Miami], Fisher’s exact test) and strong concordance of effect directions, which was greater than expected by chance (89%, p=6.56×10^−4^ [GNPC]; 78%, p=1.05×10^−3^ [Miami], one-sided exact binomial test; **Supplementary Figure 5**).

Collectively, these results demonstrate substantial overlap in dysregulated biomarkers at both pre-diagnostic and clinical ALS stages and highlight the importance of comparing biomarkers across disease stages.

### Comprehensive assessment of previously described biomarker candidates

A total of 253 proteins linked to neurodegeneration and/or ALS from various sources (see **Supplementary Methods**) were also measured in EPIC4ALS. Of these, 21 (8%) showed nominally significant associations, including TNFRSF1B (ALS KEGG pathway), TEK, CST3, KDR, TREM1, and ARSA (all on the NULISA CNS panel; **Supplementary Table 9**). Notably, none of our top pre-diagnostic biomarkers appeared among these previously described candidates. Conversely, 8 of the 27 at least suggestive pre-diagnostic ALS biomarkers overlapped with a SomaScan-based Mendelian Randomization study^19^. Of these, CA3 showed a significant causal association with ALS at FDR=0.05 (**Supplementary Table 10**).

### Cross-disease comparisons

We evaluated the 27 pre-diagnostic ALS biomarkers in incident PD (n=4,519 including 555 cases) and AD (n=1,687 including 413 cases) using Cox regression analyses in the EPIC4PD and EPIC4AD case-cohorts (**Supplementary Table 11**). None of the FDR-significant pre-diagnostic ALS biomarkers were significantly associated with pre-diagnostic PD or AD. However, the suggestive biomarkers MFAP5 and CD209 (seq.3029.52) were (proteome-wide) FDR-significant in PD, with effect estimates pointing in the same direction. No ALS biomarkers were FDR-significant for AD. Proteome-wide, 37 of 331 proteins (11%) nominally associated with ALS over 5-20 years were also nominally associated with PD or AD; while 89% were ALS-specific. No significant enrichment of PD or AD biomarkers among those identified for ALS was found (p=0.133 and p=0.134). Among proteins nominally associated with both ALS and PD or AD, 63% (PD) and 48% (AD) showed concordant directions, with a trend toward higher concordance for PD but not AD (p=0.0717 and p=0.636; exact binomial test). In summary, MFAP5 and CD209 are shared between pre-diagnostic ALS and PD, but most pre-diagnostic ALS biomarkers are disease-specific.

### Functional characterization of pre-diagnostic ALS biomarkers

Gene ontology (GO) analysis of nominally significant pre-diagnostic ALS biomarkers (p<0.05 for 5-20 year follow-up) identified nominally significant enrichment primarily in immune-related processes (hemopoiesis, innate immune response, T cell–mediated immunity, antibacterial humoral response). Additional terms included muscle structure development, proteolysis, protein secretion, cell–cell adhesion, vascular permeability, and mitochondrial catabolic processes. Triglyceride homeostasis and digestion were borderline significant. Key molecular function enrichments were for calcium ion binding, serine-type endopeptidase inhibitor activity, protein disulfide isomerase activity, and muscle-related functions. Nominally significant enrichments were also found in the extracellular region, cell surface, endoplasmic reticulum, and lysosomal lumen (**Supplementary Table 12**).

Heatmaps of RNA-seq expression data from the Human Protein Atlas [HPA] showed that several biomarkers, including CA3, KLHL41, and THAP4, were predominantly expressed in skeletal muscle (and to a lesser extent heart); other biomarkers were predominant in adipose tissue, brain (especially cerebral cortex), pancreas, and liver. Protein-level data based on immunohistochemistry (IHC) experiments available in the HPA were consistent (**Supplementary Figures 6-7**). Tissue enrichment analysis with UniProt annotations showed overrepresentation in lymphoid tissue (tonsil), pancreas, and skeletal muscle, and borderline presence in cerebrospinal fluid (**Supplementary Table 12**). Protein-protein interaction network (PPI) analysis of all FDR- and nominally significant pre-diagnostic biomarkers revealed nine functionally relevant clusters, enriching in immune-, muscle- and neuronal-related pathways (**Figure 3, Supplementary Table 13)**. The convergence of both FDR- and nominally significant ALS biomarkers in the PPI suggests that a substantial number of the nominally significant biomarkers are relevant for the disease.

**Figure 3.** Protein-protein interaction networks of pre-diagnostic ALS biomarkers. Protein-protein interaction network of all FDR-significant pre-diagnostic ALS biomarkers (larger nodes) and nominally significant pre-diagnostic ALS biomarkers (smaller nodes). The top Gene-Ontology biological process enrichment(s) for each cluster are summarised. Grey nodes clustered alone and lack enrichment.

A semi-systematic, manual literature review found prior functional evidence for involvement in ALS or its models for 21 of the 27 proteins (**Supplementary Table 14**). Finally, comprehensive interpretation with the large-language model (LLM)-based “Proteome Interpreter” suggested increased cytoskeletal remodeling, energy buffering, oxidative/nitrosative stress defense, membrane transport/excitability, and growth-factor/immune signaling, but reduced fatty acid uptake, immune sensing, protein-folding stress resolution, and extracellular matrix maintenance (**Supplementary Text**).

### Organ-specific biological aging

Next, we investigated whether incident ALS patients show signs of accelerated organ-specific aging by calculating 11 organ-specific clocks, a ‘cognition brain’ clock, as well as a non-organ-specific conventional and ‘organismal’ clock, as recently described^20^. Cox regressions on these 14 clocks revealed FDR-significant associations between incident ALS and the ‘cognition brain’ (HR=1.49, p=1.78E-03), muscle (HR=1.44, p=1.85E-03), and immune clocks (HR=1.39, p=2.51E-03), suggesting accelerated aging of incident ALS patients in these systems. Furthermore, conventional, organismal, brain, and liver clocks showed nominally significant associations (**Figure 2E, Supplementary Table 15**). Visualizing HRs in 10-year sliding windows across the complete follow-up time (0-30 years) suggested that organ-specific aging effects can be observed as early as 25-15 years before ALS-related death (20-10 years before clinical onset; **Supplementary Figure 4**). Analyses using spline regression did not reveal any significant time-varying effects (**Figure 2E, Supplementary Table 7**).

### Druggability assessment of pre-diagnostic biomarkers

Given their potential relevance early in the disease process, we assessed whether any of the 27 top ALS biomarkers are druggable using the Open Targets database. Nearly half (44%) showed evidence as potential drug targets, with CA3 being particularly prominent (**Supplementary Table 16**). The LLM model suggested therapies targeting oxidative/nitrosative stress, sarcomere stabilization, calpain inhibition, enhanced unfolded protein response, reduced fibrosis and TGF-β signaling, energy support, lipid handling, and membrane trafficking. A combined regimen was considered to be most logical including a mitochondria-targeted antioxidant, calpain inhibitor, UPR/HSP booster, with optional anti-TGF-β agent to prevent fibrosis. Early testing was recommended to comprise ROS markers (8-OHdG), calpain activity assays, ER-stress readouts (BiP, spliced XBP1), fibrosis indexes (collagen I/III) and functional outputs (contractile force, echocardiography; **Supplementary Text**).

## DISCUSSION

In this population-based, prospective case-cohort study, we present the first comprehensive characterization of ultra-early blood-based proteomic biomarker signatures for ALS. Leveraging the large European EPIC cohort with extended follow-up, high-resolution proteomic profiling, and independent indirect validation in prevalent ALS, we identified the plasma protein biomarkers SECTM1, CA3, THAP4, KLHL41, and SLC26A7 as associated with ALS up to 15-25 years before ALS-related death (or 10-20 years or more before diagnosis). All but one (SECTM1) also replicated in independent prevalent ALS cases, confirming the robustness of biomarker signals across disease stages. Overall, these findings provide important insights into the timing and underlying biology of ALS in its pre-diagnostic phase, revealing very early and sustained involvement of adaptive immune, muscle, and brain systems, all showing signs of accelerated aging and having high biological plausibility. While some pre-diagnostic protein biomarkers overlapped with PD, the ALS signature remains largely disease-specific. Thus, our study sets a new foundation for biomarker research in ALS and provides new insights that could potentially be leveraged for early detection, improved patient stratification in clinical trials, and novel therapeutic approaches.

Our study provides several key findings: First, five specific plasma proteins emerge as significant predictors of ALS many years before death/diagnosis. Our analyses of time-varying associations suggest that these markers rise well before symptoms, with the earliest associations detectable up to two decades or more years prior to diagnosis. This suggests that ALS represents a slowly evolving gradual, multi-stage molecular process rather than a quickly-evolving late-life event — mirroring the latent intervals described for PD and AD^21,22^.

Second, analysis of the temporal biomarker patterns has uncovered key pathophysiological mechanisms. GO analyses indicate that immune and muscle systems are affected early in pre-diagnostic ALS. Proteomic clocks further demonstrate that immune, muscle and brain/cognitive aging processes are pronounced very early in pre-diagnostic ALS. While there is a large body of literature supports a role for the immune system in ALS^23–25^, our data suggests that immune alterations occur at the very beginning of the pathogenesis of ALS, possibly indicating opportunities for early immune-modulating interventions. We further observed more subtle early features of metabolic processes including lipid metabolism — expanding on previous findings in presymptomatic mutation carriers^26^ and prospective studies^27,28^ — and a previously unknown, potential involvement of the digestive system (liver and pancreas).

Third, prior studies have mainly addressed candidate biomarkers in prevalent or rare familial ALS (e.g., ^29,30^). By contrast, our approach investigates the common pre-diagnostic form in a large, unselected population using large-scale proteomic profiling. Indirect validation in two independent ALS case-control datasets further supports the relevance and potential clinical utility of the identified biomarkers and illustrates that this indirect validation can help identify genuinely predictive markers. While we have not directly evaluated their predictive utility in risk modeling, the convergence of pre-diagnostic and prevalent biomarkers suggests that these markers may facilitate earlier diagnosis prior to current clinical assessment. This may also be valuable considering the substantial delays that currently often occur between initial symptom onset and clinical diagnosis.

Finally, for the first time, we applied the AI-based LLM “Proteome Interpreter” as a case study to complement our interpretation of molecular signatures and therapeutic opportunities. Notably, this tool reached conclusions consistent with our traditional functional annotation methods, efficiently mimicking expert analytical reasoning to enable rapid interpretation of high-dimensional molecular data and identification of actionable insights. Our biomarker associations — detectable up to two decades before diagnosis and stable until clinical diagnosis — open new avenues towards improvement of early diagnosis, stratification of patients in clinical trials, monitoring disease progression, and eventually timely interventions with novel disease-modifying therapies. Specifically, our findings suggest that addressing inflammation, muscle changes, and metabolic pathways at an early stage may help to favorably modify disease course and enhance outcomes for patients. Strengths of this study include the large, population-based cohort, extensive follow-up, comprehensive pre-diagnostic proteomic profiling, and independent indirect validation in two external studies. Limitations include the relatively low number of ALS cases, single pre-diagnostic blood samples, using age at death as a proxy of age at diagnosis, and potential residual confounding. Future studies should leverage repeated sampling, diverse populations, and functional approaches to clarify causal roles and translational potential of identified biomarkers.

In summary, we redefine ALS as a condition with a prolonged molecular pre-diagnostic phase — characterized by immune, muscular, metabolic, and brain involvement detectable in blood proteomes. Our results substantially advance the understanding of ALS pathophysiology and underscore new prospects for biomarker-driven early detection and intervention. Strategic application of blood-based proteomics could ultimately improve ALS diagnosis, care, and prevention.

## Data Availability

Data can be accessed by external researchers after approval by the EPIC4ND working group and the EPIC steering committee. Potential collaborators can contact the EPIC4ND working group chair, Christina Lill and the EPIC administrator Sherry Morris.

## Acknowledgements

The authors thank all EPIC participants and the countless scientists who have contributed to the EPIC cohort over the past 30 years. We thank the National Institute for Public Health and the Environment (RIVM), Bilthoven, the Netherlands, for contributing cases and ongoing support to the EPIC study. We sincerely thank Bertrand Hemon for his assistance with data management, and Prof. Paolo Vineis, Prof. Martin Wolkewitz, and Dr. Jürgen Wellmann for helpful discussions. This work was supported by NIHR Imperial Biomedical Research Centre at Imperial.

## Disclaimer

Where authors are identified as personnel of the International Agency for Research on Cancer / World Health Organization, the authors alone are responsible for the views expressed in this article and they do not necessarily represent the decisions, policy or views of the International Agency for Research on Cancer / World Health Organization.

## Authors’ contributions

Study concept and supervision: ER, MJG, PF, CML; acquisition/contribution of data: JMH, MG, AJZ, MJS, CTS, SMCY, SA, DP, SS, SP, TJK, NW, RK, RCT, RV, GNPC, CS, GM, VK, LM, ER,

MJG, PF, CML, data analysis and interpretation: JH, RKL, VV, SM, VD, LD, JS, KSB, DB, YZ, JW, FA, FH, OO, ML, MK, IT, AE, MM, MB, LB, ER, MJG, PF, OR, CML; drafting the manuscript: JH, CML. Critical revision of the manuscript for content: all co-authors

## METHODS

A full description of the methods employed in this study is provided in the Supplementary Methods.

### Discovery cohort and study design

The EPIC4ALS case-cohort analyzed here is embedded within EPIC, a prospective study of 519,978 participants enrolled between 1991 and 2000 across 10 European countries, and aims to identify molecular biomarkers prior to the emergence of clinically manifest ^16^. ALS case ascertainment was based on a source population of 222,613 subjects from 12 centers across five countries (Spain, Italy, Netherlands, UK, and Germany), as described previously^16^. Given the fatal nature and short survival after diagnosis^4^, incident cases were identified via mortality records as previously described^16^. To identify pre-diagnostic ALS biomarkers, a 5-year wash-out phase between blood draw and age at death/censoring was applied, excluding 21 ALS patients and 149 subcohort members from most analyses.

At baseline, data on diet, lifestyle, medical history, and anthropometric measurements were collected. Citrate plasma samples were processed using standardized protocols and stored in liquid nitrogen at the EPIC biobank at IARC in Lyon, France. Participants are followed by regular registry linkage and/or questionnaires. The study received ethics approval from IARC and all relevant boards; all participants provided written informed consent.

### Generation and quality control of proteomic data in EPIC4ALS

Protein abundances were quantified in plasma samples using the SomaScan 7K (v4.1) platform (SomaLogic, Boulder, CO, USA), which employs chemically modified DNA aptamers for high-throughput, high-specificity protein detection^32^. The 7K version quantifies 7,596 aptamers targeting 6,432 unique proteins^33^. Proteomic profiling was conducted for 17,841 EPIC participants selected for various endpoints at SomaLogic, using additional control probes for quality control (QC). Sample and data processing are detailed in the **Supplementary Methods**.

After normalization and QC, aptamer measurements were log10- and z-transformed to mean=0, SD=1 in the subcohort. Values deviating more than 5 SD from the mean were ‘capped’ at 5 SD. For all significant findings, log_10_-transformed values were visually inspected (**Supplementary Figure 7**).

### Cox proportional hazard regression analyses in EPIC4ALS

The final effective EPIC4ALS case-cohort for Cox proportional hazard analyses included 4,567 participants (172 incident ALS cases) with 7,285 aptamers (6,381 proteins) for most analyses (i.e., those applying the 5-year wash-out period). We used Prentice-weighted^34^ Cox regression models to examine associations between aptamers and risk of ALS-related death, which we used as a proxy indicator for ALS diagnosis, since clinical diagnosis data were not available for all participants. Age was used as a time scale^34^. Age at death served as a proxy for diagnosis. The basic model was stratified by 5-year age intervals, sex, and center, and a 5-year wash-out was applied to exclude prevalent cases (and the corresponding non-cases) dying within the first 5 years. Analyses were performed for both full (5-30 years) and restricted (5-20 years) follow-up. Proteomic associations were considered significant at FDR 0.05. The proportional hazard assumption for the top biomarkers was tested by calculating Schoenfeld residuals (using the cox.zph() function). For the top biomarkers, we also performed sex-specific analyses and tested sex effects by interaction analyses^35^.

In sensitivity analyses for the top biomarkers, we tested a full adjustment model including the baseline covariates ‘education’, ‘smoking status’, ‘BMI’, ‘physical activity’, ‘postmenopausal status’, and ‘hormone use’. We also repeated analyses excluding protein outlier values (>5 SD from the mean), acknowledging that outliers may either signal true ALS biology or unrelated technical/biological variation.

### Assessment of time-varying effects of ALS-associated proteins

Time-varying effects of ALS-associated proteins were explored using different approaches across the entire EPIC4ALS dataset with 0-30 year follow-up, i.e., using 4,737 EPIC participants including 193 ALS cases (i.e., re-including the 21 incident ALS patients and 149 non-cases who died or were censored within five years of baseline). To visualize effect estimates of top results in different time windows, HRs were computed and plotted using 10-year sliding windows in one-year increments. Exploratory analyses of time-varying effects of ALS-associated proteins were tested analyzing Cox models (Prentice-weighted, adjusting for age and stratified for sex, and center) with a ‘protein level × log time-to-event’ interaction term, and by fitting Cox models incorporating natural splines to assess non-linear time-dependent effects. For all these analyses, we used time since blood collection instead of age as the time scale. For the splines, we split the follow-up time into fine intervals (using the survSplit() function) and applied a time-varying coefficient (TVC) approach for each protein using spline interactions (protein × spline(time-to-event)). The Prentice-weighted Cox models were used adjusting for age (as a time-varying variable) and stratifying by sex and center. The nsk() function from the R package splines was used with 2 degrees of freedom. We evaluated the statistical evidence for time-varying effects using Wald tests.

### Indirect validation and characterization of pre-diagnostic EPIC4ALS biomarkers in clinical datasets

FDR-significant pre-diagnostic biomarkers from EPIC4ALS were evaluated for association with prevalent ALS using a large SomaScan 7K profiled case-control dataset (260 ALS, 689 controls) from the GNPC initiative (https://www.neuroproteome.org/). Only datasets with both cases and controls were included. Control participants were those without a documented clinical diagnosis of ALS, mild cognitive impairment, dementia, or PD. Protein data were log10-transformed, standardized, and outliers (>5 SD) excluded. Residuals were obtained by removing age, sex, and center effects (removeBatchEffect, limma), on which the first two principal components (PC) from PC analysis were calculated. Logistic regression adjusted for age, sex, center, and PCs was performed, along with sex-stratified analyses (adjusted for age and PCs) for biomarkers with sex-specific effects in EPIC4ALS. Additionally, summary statistics from the Miami SomaScan 7K dataset (157 ALS cases, 163 controls)^31^ were used. To assess the enrichment of clinical biomarkers among pre-diagnostic biomarkers, we conducted a one-sided Fisher’s exact test, testing whether FDR-significant clinical biomarkers were overrepresented among all pre-diagnostic biomarkers that reached nominal significance. Concordance of effect estimates in between pre-diagnostic and clinical datasets was tested using a one-sided binomial exact test.

### Comparison to previous studies

We compiled a list of 520 unique candidate biomarkers previously linked to neurodegeneration and/or ALS based on a range of different resources (**see Supplementary Methods**). The 253 proteins from this list measured in EPIC4ALS were tested for association and controlled using FDR=0.05.

### Cross-disease comparisons

For cross-disease comparisons, we analyzed FDR-significant pre-diagnostic ALS biomarkers in pre-diagnostic AD and PD using Prentice-weighted Cox regression of SomaScan 7K data in the EPIC4AD case-cohort (1,687 participants, including 413 incident AD cases, 2-20 years follow-up) and EPIC4PD (4,519 participants, 555 PD cases, 2-20 years; for a description of the case-cohorts, see ref. ^16^). Analyses were stratified by 5-year age groups, sex, and center. Using a one-sided Fisher’s exact test, we assessed whether significant associations in AD and PD were enriched among proteins nominally significant in ALS. A one-sided binomial test evaluated if overlapping nominally significant proteins had concordant effect directions more often than expected by chance.

### Expression and protein abundances of top biomarkers

Gene expression and protein abundance of top ALS biomarkers across tissues were assessed using bulk RNA-Seq and IHC data from the HPA (v24.0, Ensembl 109, accessed May 6th, 2025)^36^. Pre-diagnostic proteins were annotated using ‘Consensus RNA data’ for 50 tissues and IHC data for 45 tissues. Heatmaps were generated using hierarchical clustering and gene-scaled expression values to highlight tissue-specific variation.

### Gene set and cell type enrichment analyses

Gene set and tissue enrichment was performed using DAVID (v2024q4; https://davidbioinformatics.nih.gov/home.jsp). All unique proteins nominally associated with incident ALS were tested for enrichment in ‘biological process’, ‘molecular function’, and ‘cellular component’ GO categories, as well as UniProt Tissue Annotations. The statistical overrepresentation tests were conducted with the background set to all analyzed SomaScan proteins. To minimize spurious results, only results based on at least three pre-diagnostic proteins were considered. Analyses were controlled at FDR=0.05.

### Protein-protein interaction network analysis

Protein-protein interaction networks were generated using the STRING database (v12.0) for all FDR-significant and nominally significant EPIC4ALS biomarkers and visualized with igraph (v2.1.4). Interactions with a minimum interaction score of 0.4 were based on the following sources: ‘textmining’, ‘experiments’, ‘databases’, ‘co-expression”, “neighborhood’, ‘gene fusion’, and ‘co-occurrence’. The network was clustered using a fast greedy algorithm, top GO enrichments for each cluster examined, and unconnected nodes removed.

### Organ-specific biological aging

Organ-specific biological age estimates were calculated using the organage package^20^. In this context, organ age gaps represent organ-specific age acceleration measures by comparing chronological and estimated biological age based on specific sets of organ-specific proteins. Organ age gaps (z-scored) served as exposure variables in Cox proportional hazard models (with Prentice weights and stratified by 5-year age group, sex, and center) to assess associations with ALS incidence. While Oh et al. described the ‘cognition’ ages for all organs^20^, we only analyzed the ‘cognition brain’ age. We applied FDR control to account for the 14 organ age estimates analyzed.

### Drug Repurposing

We used the Open Targets platform^37^ to assess druggability and repurposing potential for all FDR-significant, putative pre-diagnostic ALS biomarkers. Focusing on small-molecule evidence only, we screened ∼17,000 small-molecule drugs and 21,087 protein targets for interactions with the top 27 ALS biomarkers. Druggability was defined as reported direct binding or predicted high-quality pharmacological modulation, based on curated experimental, structural, or clinical Open Targets data.

### Large language model

The data was further analyzed using a proteomics-specific large language model (LLM)-based tool designed to expedite proteomic data interpretation (developed by the Mann laboratory, https://github.com/MannLabs/alphapeptstats/releases/tag/v0.7.1). This tool integrates the generalized abilities of LLMs with domain-specific resources and real-time knowledge retrieval. The primary input consisted of the list of all analyzed proteins including effect direction and p value (from the 5-20 year follow-up). For each protein, information such as name, gene symbol, and functional description was automatically obtained from UniProt to provide curated context to the LLM. This information was combined with a pre-defined prompt specifying persona, style, and output format, as well as a chain-of-thought (CoT) instruction, mirroring the reasoning process of a human scientist. The complete prompt was submitted to the OpenAI o3 API, which had demonstrated the best internal performance for this task, to achieve a comprehensive interpretation of the molecular ALS signature. The analysis was performed in three replicates to assess the consistency of responses. Additionally, a targeted follow-up query regarding potential therapeutic opportunities was included to identify initial avenues for exploring clinical interventions.

